# Diagnostic performance of multiplex lateral flow tests in ambulatory patients with acute respiratory illness

**DOI:** 10.1101/2024.03.18.24304455

**Authors:** Caitriona Murphy, Loretta Mak, Samuel M. S. Cheng, Gigi Y. Z. Liu, Alan M. C. Chun, Katy K. Y. Leung, Natalie Y. W. Sum, Eero Poukka, Malik Peiris, Benjamin J. Cowling

## Abstract

**Background:** We assessed the performance of three different multiplex lateral flow assays which provide results for influenza, respiratory syncytial virus (RSV) and SARS-CoV-2.

**Methods:** Ambulatory patients 6 months and older presenting with two or more symptoms or signs of an acute respiratory illness were enrolled in an outpatient clinic in Hong Kong. Multiplex lateral flow tests manufactured by SureScreen, Microprofit and Goldsite were performed by trained research staff using the nasal swabs from each test kit, and separate swabs were collected for RT-PCR testing.

**Results:** Between 4 April and 20 October 2023, 1646 patients were enrolled and tested by at least one lateral flow test. The point estimates for all three multiplex tests had high sensitivity above 80% for influenza A and SARS-CoV-2, and the tests manufactured by Microprofit and Goldsite had sensitivity exceeding 84% to detect RSV. Test sensitivity increased with viral load. Specificity was higher than 97% for all three tests except for the SureScreen test which had specificity 86.2% (95% CI: 83.9% to 88.3%) for influenza A.

**Conclusions:** The multiplex lateral flow tests provided timely diagnosis of influenza, RSV and SARS-CoV-2 infection and can be used to inform clinical management and infection control such as isolation behaviours.

## INTRODUCTION

Timely and accurate diagnosis of infection from respiratory viruses is essential for managing clinical care and reducing transmission. Lateral flow tests, also referred to as rapid antigen tests, are point of care devices that can identify the presence of an infectious disease by detecting microbial proteins within 30 minutes. The key advantage of lateral flow tests over the current gold standard, polymerase chain reaction (PCR) assay is the lower cost and the quick turnaround time for results. Additionally, performing PCR tests requires trained staff and laboratory infrastructure that is resource intensive and may not always be available. Lateral flow tests were widely distributed during the COVID-19 pandemic to improve diagnosis of cases in the community and facilitate more efficient isolation and quarantine policies. The use of lateral flow tests were deemed a successful strategy in multiple locations for use in the community [1-3] and subgroups of the community where repeated testing using lateral flow tests was utilised to help keep schools and workplaces open [4-7].

In some locations, lateral flow tests are now available for detecting influenza A/B, SARS-CoV-2 and RSV in a single test kit. Diagnostic tests that can quickly identify the presence of infection while also indicating which respiratory virus an individual is infected with will aid patient management, particularly when specific antivirals should be administered rapidly after symptom onset. The turnaround time of lateral flow tests allows patients to receive appropriate antiviral treatment in a timely manner and when it may be most beneficial during the course of infection. The objective of this study was to assess the performance of three different multiplex lateral flow tests for detecting SAR-CoV-2, influenza A/B and RSV compared to RT-PCR in an outpatient setting in Hong Kong.

## METHODS

### Study participants

This study included outpatients enrolled in an ongoing influenza vaccine effectiveness study in Hong Kong. Patients at least 6 months of age were enrolled if they presented with acute respiratory illness defined as having at least two of seven signs/symptoms (fever ≥37.8°C, cough, sore throat, runny nose, headache, myalgia and phlegm) within 72 hours of symptoms onset. Participants or their legal guardians were provided with a questionnaire to obtain demographic details as well as vaccination history.

### Rapid test procedures

Upon obtaining informed consent, two lateral flow assays namely “Fluorecare SARS-CoV-2 & Influenza A/B & RSV Antigen Combo Test Kit (Self-Test)” (Shenzhen Microprofit Biotech Co. Ltd., Shenzhen, China) and “SARS-CoV-2 + Flu A&B Antigen Combo Rapid Test Cassette (Nasal Swab)” (SureScreen Diagnostics Ltd, Derby, UK) were administered by the research team. If either of these tests were unavailable at the time of participant enrolment a third test, “SARS-CoV-2 & Influenza A/B & RSV Antigen Kit” (Colloidal Gold) (Goldsite Diagnostics Inc., Shenzhen, China) was used. Throughout the paper, the three multiplex lateral flow tests are named after their manufacturer, Microprofit, SureScreen and Goldsite respectively.

The Microprofit and Goldsite tests are combination tests for detecting SARS-CoV-2, influenza A/B and RSV, while SureScreen detects SARS-CoV-2 and influenza A/B. All three tests use a nasal swab and provide results within 15 minutes. Further details of the test specifications are available in the Supplementary (Supp. Table 1; Figures 1 to 3). In the event that a lateral flow tests result was invalid (i.e. there was no visible coloured band for the control line) a second test was conducted but not a third test if the second was also invalid. Those that had a third invalid test result were removed from the analysis.

**Table 1:** The performance of lateral flow tests for detecting influenza A, SARS-CoV-2 and RSV infection.

| Respiratory virus and test manufacturer | Sensitivity |  | Specificity |  |
| --- | --- | --- | --- | --- |
|  | % | 95% CI | % | 95% CI |
| **Influenza A** |  |  |  |  |
| Microprofit | 82.1 | 77.0 to 86.5 | 97.6 | 95.9 to 98.7 |
| Goldsite | 84.9 | 80.9 to 88.3 | 98.0 | 96.2 to 99.1 |
| SureScreen | 89.7 | 87.0 to 91.9 | 86.2 | 83.9 to 88.3 |
| **SARS-CoV-2** |  |  |  |  |
| Microprofit | 81.7 | 73.5 to 88.3 | 99.4 | 98.5 to 99.8 |
| Goldsite | 80.4 | 67.6 to 89.8 | 99.4 | 98.5 to 99.8 |
| SureScreen | 88.2 | 82.3 to 92.6 | 98.1 | 97.2 to 98.7 |
| **RSV\*** |  |  |  |  |
| Microprofit | 94.6 | 81.8 to 99.3 | 97.6 | 94.1 to 99.4 |
| Goldsite | 84.2 | 68.7 to 94.0 | 99.5 | 97.1 to 100 |

\*The SureScreen test did not include RSV

**Figure 1.**
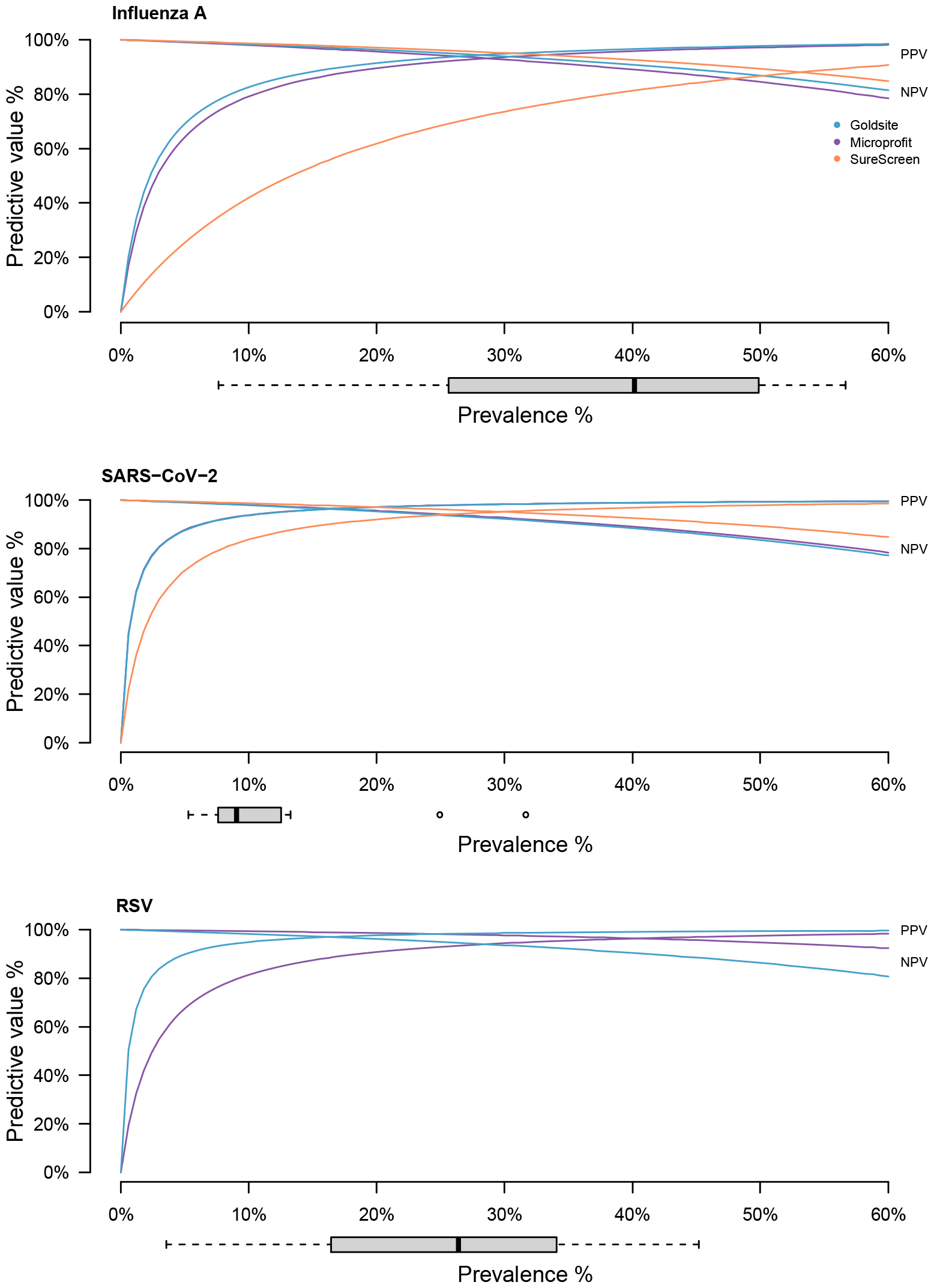
Relationship between the positive and negative predictive values and prevalence for each test. Predictive values were estimated using the sensitivity and specificity presented in Table 1. The specificity for Microprofit and Goldsite were the same for detecting SARS-CoV-2 and the SureScreen test did not include RSV. The shaded areas represent the 95% confidence intervals and the prevalence of each respiratory virus, during the study period, are represented by the grey boxplots.

### Laboratory testing

A separate set of nose and throat swabs were also collected by the research team or attending physician and were pooled into a single vial of transport medium and transported to the laboratory for testing. All samples were tested by reverse transcription-polymerase chain reaction (RT-PCR) for influenza (A and B) and SARS-CoV-2. Moreover, all lateral flow test RSV-positive and a randomly selected subset of lateral flow test RSV-negative samples were tested for RSV using RT-PCR.

A standard 20□ µL RT-PCR assay was performed, comprising 5 □ μL of a 4X master reaction mixture (TaqMan Fast Virus 1-Step Master Mix, ThermoFisher), 0.5□ µM of the forward primer, 0.5□ µM of the reverse primer, 0.25□ µM of the probe, and 2□ μL of the RNA sample. The RT-PCR reactions were carried out using a ViiA7 Real-Time PCR system (ThermoFisher) with the following thermal cycling conditions: reverse transcription at 50°C for 5□ minutes, inactivation of reverse transcriptase at 95°C for 20□ seconds, followed by 40 cycles of PCR amplification (denaturation at 95°C for 5□ seconds, annealing/extension at 58°C for Influenza A and B, SARS-CoV-2, 50°C for RSV A and B for 30□ seconds). Primer and probe sequences can be found in Supplementary Table 2. The Ct-value was evaluated from all the PCR positive samples. A false-positive result was defined as a positive result for a particular virus on a lateral flow test and a subsequent negative confirmatory RT-PCR result for that virus.

### Statistical analysis

The sensitivity and specificity of each of the lateral flow tests was estimated using RT-PCR confirmed infection for each virus as the comparator. The incidence of influenza B was low during the study period and therefore, the performance of lateral flow tests for detecting influenza B was not evaluated. Multivariable logistic regression, including the RT-PCR positives was used to evaluate the relationship between lateral flow test positivity and age, sex, vaccination status and symptom onset. Positive and negative predictive values (PPV and NPV) were estimated conditional on different values for the true prevalence. Confidence intervals (CI) were estimated using binomial distributions. All statistical analyses were conducted using R version 4.2.2 (R Foundation for Statistical Computing, Vienna, Austria).

### Ethical approval

The study protocol was approved by the Institutional Review Board of the University of Hong Kong. The lateral flow tests aided with clinical management, and written informed consent was obtained for each participant and parental consent was obtained for participants below 18 years of age.

## RESULTS

Between 4 April and 20 October 2023, 1646 outpatients with acute respiratory symptoms were enrolled. Males accounted for 47.0% and 63.3% were children (<18 years old). The majority (26.3%) of adults were less than 50 years old with just 4.1% being 65 years of age or older. All patients were tested for influenza and SARS-CoV-2 by RT-PCR. Influenza A was detected in 651 (39.6%) patients and 171 (10.4%) were laboratory confirmed SARS-CoV-2. During our study period the predominant SARS-CoV-2 strains in the community were Omicron XBB subvariants. Of the 431 samples tested for RSV by RT-PCR 75 (17.4%) were positive. The Microprofit test was used to test 814 (49.5%) patients, and 832 (50.5%) were tested using the Goldsite test. SureScreen was performed by 1632 (99.1%) patients and the reasons for not being tested for the remaining 23 patients were because the test was out of stock (7/23), they were not feeling well (6/23), were too busy (3/23) or other unspecified reasons (7/23).

The SureScreen test had a significantly higher sensitivity (89.7%, 95% CI: 87.0% to 91.9%) compared to Microprofit (82.1%, 95% CI: 77.0% to 86.5%) and Goldsite (84.9%, 95% CI: 80.9% to 88.3%) for detecting influenza A but performed similarly for detecting SARS-CoV-2. Both the Microprofit and Goldsite tests were comparable for detecting RSV (Table 1). This is reflected in the NPV estimates for different values of prevalence in Figure 1. When the viral load was high (i.e. low CT value) all three tests had a higher sensitivity compared to patients with low viral loads particularly for detecting influenza A and SARS-CoV-2. The sensitivity ranged from 87.5% to 100% for detecting CT values <25 and from 28.6% to 75.0% for detecting CT values ≥30 (Figure 2). Microprofit and SureScreen had a significantly lower sensitivity among those 65 years and older compared to younger age groups for detecting influenza A (Supp. Table 2; Figure 4). Sensitivity for those 65 years and older was 25.0% (95% CI: 3.2% to 65.1%) and 70.8% (95% CI: 48.9% to 87.4%) compared to 86.3% (95% CI: 73.7% to 94.3%) and 94.6% (95% CI: 88.7% to 98.0%) for those 6 months to 5 years old for Microprofit and SureScreen respectively. Lateral flow tests were more sensitive for detecting influenza A in patients with a symptom onset between 1 to 2 days prior to presentation (odds ratio: 1.2, 95% CI: 1.1 to 4.7) compared to those with symptom onset within 1 day. The estimates of test sensitivity for patients that were vaccinated for influenza and COVID-19 were similar to those who had not been vaccinated for the respective pathogen (Supp. Table 3).

**Figure 2.**
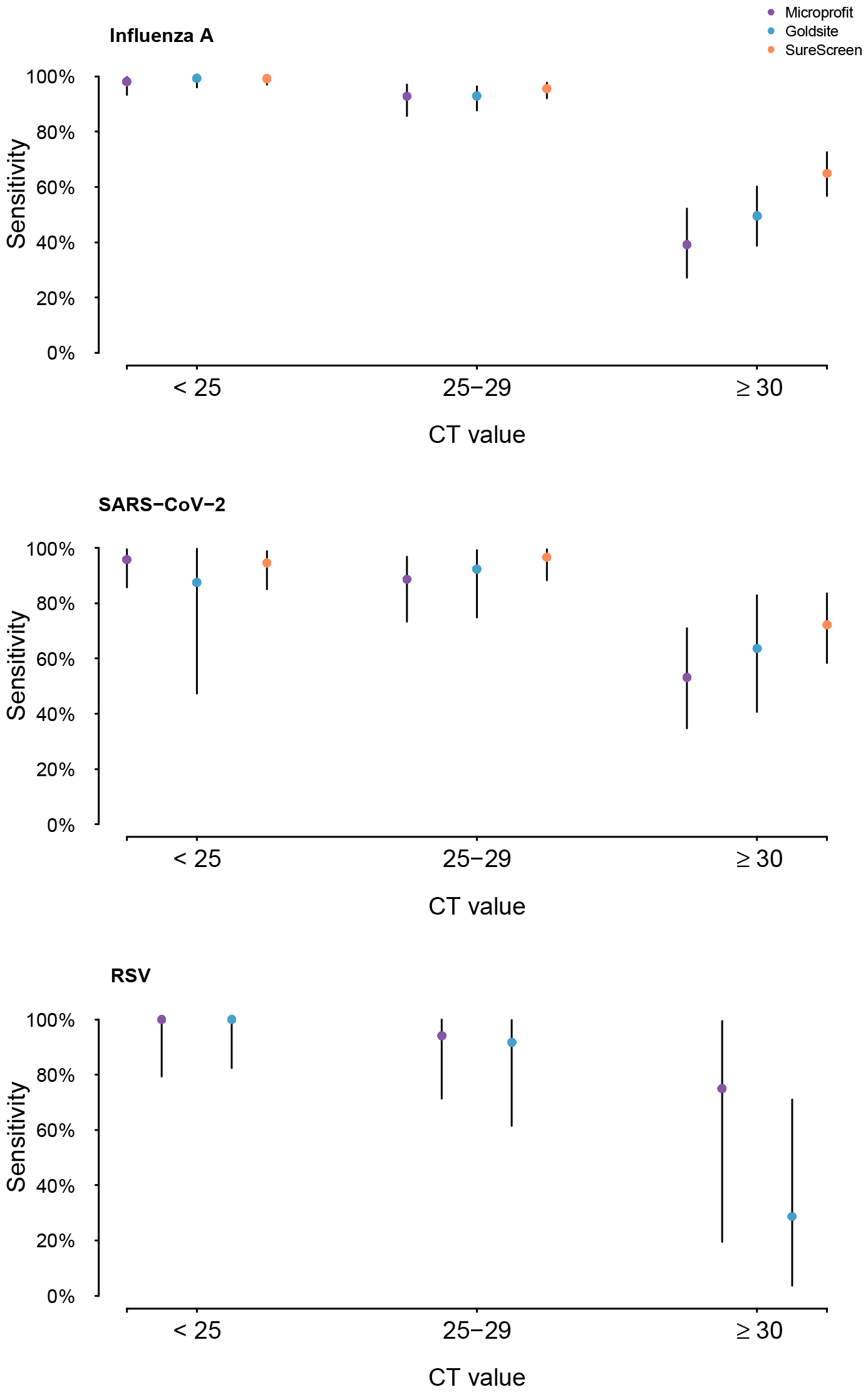
Sensitivity with 95% confidence intervals of each lateral flow test by Ct value category.

Specificity for the Microprofit and Goldsite tests were significantly higher than SureScreen for detecting both influenza A and SARS-CoV-2, resulting in a lower PPV for SureScreen (Figure 1). Other than the lower specificity for SureScreen for detecting influenza A (86.2%, 95% CI: 83.9% to 88.3%), the remaining test and virus specific point estimates had a specificity over 97% (Table 1). Specificity was similar across age groups and vaccination status. Among the 138 influenza A false positives 8 (5.8%) were laboratory confirmed SARS-CoV-2 infections and among the SARS-CoV-2 false positives (n=30) and RSV false positives (n=5) there were 3 patients among each that tested positive for influenza A by RT-PCR (Supp. Table 4).

## DISCUSSION

We evaluated the performance of three multiplex rapid tests, Microprofit, Goldsite and SureScreen in symptomatic individuals in an ambulatory care setting in Hong Kong compared to the gold standard of RT-PCR. Point estimates of Microprofit and Goldsite tests reached the WHO minimum test performance for all three viruses with a sensitivity of ≥80% and specificity ≥97% [8]. SureScreen did not meet these requirements for all targeted respiratory viruses in a single test kit. Instead it had a high sensitivity for detecting influenza A and SARS-CoV-2 but had a lower specificity for detecting influenza A (86.2%, 95% CI: 83.9% to 88.3%). The study period was during the seasonal influenza epidemic in Hong Kong, with one epidemic peak in late April dominated by influenza A(H1N1) and a second peak in September dominated by influenza A(H3N2) [9]. An epidemic of SARS-CoV-2, dominated by Omicron XBB subvariants also occurred from April to July 2023 [9] and therefore, we expected the resulting PPV and NPV during periods of higher prevalence to be moderate to high.

The estimates in this study are higher than those observed in some previous studies [10-15]. Although, studies estimating the performance of the same tests as those evaluated here were among asymptomatic individuals and we were unable to find studies using the same multiplex tests among symptomatic individuals. The varying sensitivity for symptomatic and asymptomatic individuals has been documented [16] but a systematic review also identified that the performance of lateral flow tests in symptomatic individuals had a wide range (34.3% to 91.3%) depending on the manufacturer [16]. We identified one study estimating the performance of a multiplex Microprofit test that reported a sensitivity for detecting influenza A of 80.8% (95% CI: 67.2% to 94.4%) among asymptomatic individuals. However, the test sensitivity was 41.5% (95% CI: 26.2% to 56.8%) for detecting RSV in that study [12] compared to 94.6% (95% CI: 81.8% to 99.3%) estimated in this study. The performance of a singleplex SureScreen test for detecting SARS-CoV-2 was evaluated in inpatients and outpatients from a single centre in the UK with a lower estimated sensitivity of 65% (95% CI: 55.2% to 73.6%) but a very high specificity at 100% (95% CI: 96.3% to 100%) [10]. Another study using a singleplex SureScreen diagnostic test to detect SARS-CoV-2 estimated a sensitivity and specificity of 28.8% (95 CI%: 20.2% to 38.6%) and 97.8% (95% CI: 94.5% to 99.4%) respectively for mass screening in unexposed asymptomatic individuals [11]. Compared to the sensitivity and specificity estimates reported by the manufacturers (Appendix Table 1), our study estimates, in a real-world context, were slightly lower.

The estimated sensitivity of the three lateral flow tests increased as the viral load increased which is consistent with previous literature [17-20]. Some studies suggest that the viral load is higher in patients with more severe outcomes, especially considering severe cases might take longer to clear infection [21]. In turn, if the sensitivity is higher for detecting severe cases, the use of lateral flow tests could support timely prescription of antivirals. A lower sensitivity for those with a lower viral load could have negative implications when implementing lateral flow tests. Of the 88 false negatives in this study, 76% had lower viral loads. Falsely testing negative may lead to a failure to isolate and a missed opportunity to reduce onwards transmission, albeit this typically occurred among individuals with lower Ct values who might perhaps be less contagious. Health authorities in Hong Kong currently recommend symptomatic individuals use a lateral flow test and for those who test negative it is recommended to remain cautious and repeat the test over a few days [22].

Our evaluation of lateral flow test performance has limitations. While RT-PCR is often considered a gold standard it can detect inactive virions for a few weeks after infection [23]. An overestimate of infections identified by RT-PCR may subsequently underestimate the sensitivity of lateral flow tests if the objective is to identify contagious individuals and individuals early in their course of disease when antivirals would be more effective and transmission-reducing measures such as isolation could be implemented more effectively. Data were collected prospectively based on the presence of symptoms and with symptom onset occurring within three days and therefore there was a smaller sample of patients with Ct values >30, particularly for RSV. Viral culture was not carried out in this study to confirm the relationship between infectiousness and testing positive by lateral flow tests. Finally, the sensitivity may vary depending on the strains of SARS-CoV-2 circulating which in our study period included Omicron XBB subvariants. The SureScreen test evaluated here target the more conserved nucleocapsid protein of SARS-CoV-2 (as opposed to the spike protein), however variation in the test performance may persist across virus lineages [14]. It was unclear which SARS-CoV-2 protein Microprofit and Goldsite targeted.

To conclude, each of the three evaluated multiplex tests performed well in detecting one of the viral antigens in the multiplex lateral flow test but necessarily all targeted respiratory viruses. The use of lateral flow tests has been more limited in healthcare settings where PCR is readily available. However, they can efficiently implement control measures and providing an accurate diagnosis will guide appropriate treatment.

## Data Availability

All data produced in the present work are contained in the manuscript

## ACKNOWLEDGMENTS

The authors thank Julie Au for technical support. This research was supported by the Health and Medical Research Fund, Health Bureau, The Government of the Hong Kong Special Administrative Region (grant no. INF-HKU-3). BJC is supported by the Theme-based Research Scheme (Project No. T11-712/19-N) of the Research Grants Council of the Hong Kong SAR Government.

