## Appendix for "Diagnostic performance of multiplex lateral flow tests in ambulatory patients with acute respiratory illness"

Appendix Table 1: Detailed specifications of the three lateral flow tests used in our study.

| **Assay** | fluorecare® SARS-CoV-2 & Influenza A/B & RSV Antigen Combo Test Kit (Self-Test) | SARS-CoV-2 & Influenza A/B & RSV Antigen Kit (Colloidal Gold) | SARS-CoV-2 + Flu A&B Antigen Combo Rapid Test Cassette (Nasal Swab) |
|---|---|---|---|
| **Manufacturer** | Shenzhen Microprofit Biotech Co. Ltd., Shenzhen, China | Goldsite Diagnostics Inc., Shenzhen, China | SureScreen Diagnostics Ltd, Derby, UK |
| **Country of Origin** | P.R. China | P.R. China | UK |
| **Certification** | CE-IVD | CE-IVD | CE-IVD |
| **Swab type** | Nasal swab | Nasal swab | Nasal swab |
| **Reported sensitivity and specificity** | Sensitivity / Specificity<br>SARS-CoV-2: 92.9% / 100%<br>Influenza A: 92.0% / 100%<br>Influenza B: 90.9% / 100%<br>RSV: 95.5% / 100% | Sensitivity / Specificity<br>SARS-CoV-2: 93.0% / 100%<br>Influenza A: 85.0% / 99.3%<br>Influenza B: 95.0% / 100%<br>RSV: 92.3% / 100% | Sensitivity / Specificity<br>SARS-CoV-2: 99.2% / 99.5%<br>Influenza A: 90.0% / 96.8%<br>Influenza B: 93.6% / 98.2% |
| **Format** | Card | Cassette | Cassette |
| **Method** | Colloidal Gold | Colloidal Gold | Colloidal Gold |
| **Volume applied into card/cassette** | 2 drops (~60μL) | 2 drops (~60μL) | 3 drops (~80μL) |
| **Incubation** | 15 minutes | 15 minutes | 10 minutes |
| **Readout** | Visual: coloured band | Visual: coloured band | Visual: coloured band |
| **Limit of detection** | SARS-CoV-2: 49 $TCID_{50}$/mL<br>Influenza A(H3N2): 4.0 x $10^4$ $TCID_{50}$/mL | SARS-CoV-2: 49 $TCID_{50}$/mL<br>Influenza A(H3N2): 4.0 x $10^4$ $TCID_{50}$/mL | SARS-CoV-2: 1000 $TCID_{50}$/mL |

| | | | |
|---|---|---|---|
| | Influenza A(H1N1): 2.0 x $10^4$ TCID $_{50/}$mL<br>RSV A:1.2 x$10^4$ TCID $_{50/}$mL<br>RSV B : 1.6 x $10^4$ TCID $_{50/}$mL | Influenza A(H1N1): 2.0 x $10^4$ TCID $_{50/}$mL<br>RSV: 1.6x10^4 TCID $_{50/}$mL | |
| **Cross reactivity against other human virus** | The results showed no cross reactivity | The results showed no cross reactivity | The results showed no cross reactivity |

Appendix Table 2: Primers and probes for TaqMan amplification of viral RNA from influenza A, influenza B, SARS-CoV-2, RSV A and RSV B.

| Target gene | Target | Primer/ Probe | Sequence (5′-3′) | Reference |
|---|---|---|---|---|
| Influenza A | M gene | Forward | CTTCTAACCGAGGTCGAAACGTA | Terrier, Olivier et al. 2011[1] |
| | | Reverse | GGTGACARGATTGGTCTTGTCTTTA | |
| | | Probe | TCAGGCCCCCTCAAAGCCGAG | |
| Influenza B | M gene | Forward | CACAATTGCCTACCTGCTTTCA | Lambert, Stephen B et al. 2008 [2] |
| | | Reverse | GCATCTTTTGTTTTTTATCCATTC | |
| | | Probe | GCTAGTTCTGCTTTGCCTTCTCCATCTTCT | |
| SARS-CoV-2 | nsp14 | Forward | TGGGGYTTTACRGGTAACCT | Chu, Daniel K W et al. 2020 [3] |
| | | Reverse | AACRCGCTTAACAAAGCACTC | |
| | | Probe | TAGTTGTGATGCWATCATGACTAG | |
| | N gene | Forward | TAATCAGACAAGGAACTGATTA | |
| | | Reverse | CGAAGGTGTGACTTCCATG | |
| | | Probe | GCAAATTGTGCAATTTGCGG | |
| RSV A | N gene | Forward | AGATCAACTTCTGTCATCCAGCAA | van Elden, L J R et al. 2003 [4] |
| | | Reverse | TTCTGCACATCATAATTAGGAGTATCAAT | |
| | | Probe | CACCATCCAACGGAGCACAGGAGAT | |
| RSV B | N gene | Forward | AAGATGCAAATCATAAATTCACAGGA | |
| | | Reverse | TGATATCCAGCATCTTTAAGTATCTTTATAGTG | |
| | | Probe | TTCCCTTCCTAACCTGGACATAGCATATAACATACCT | |

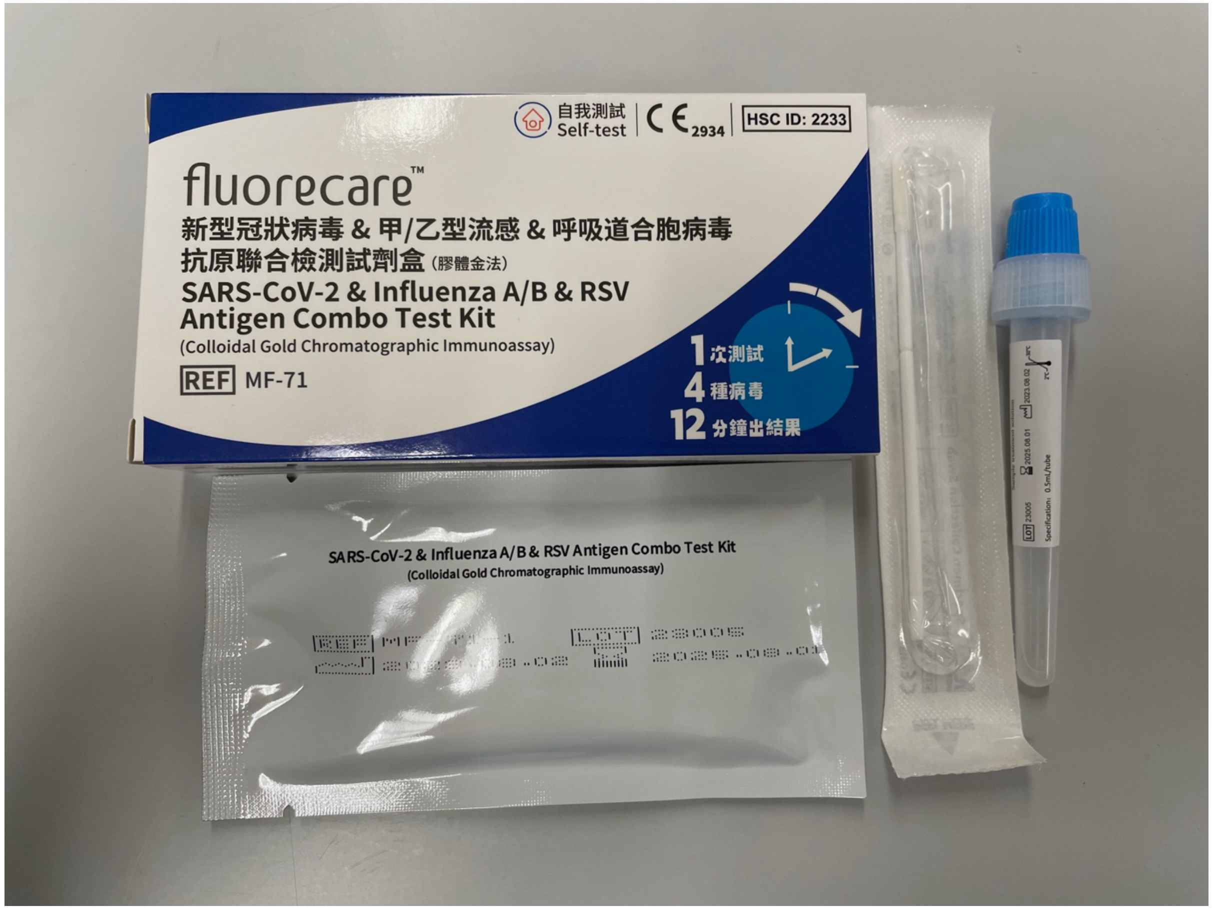

Appendix Figure 1: fluorecare SARS-CoV-2 & Influenza A/B & RSV Antigen Combo Test Kit (Self-Test) (Shenzhen Microprofit Biotech Co. Ltd., Shenzhen, China)

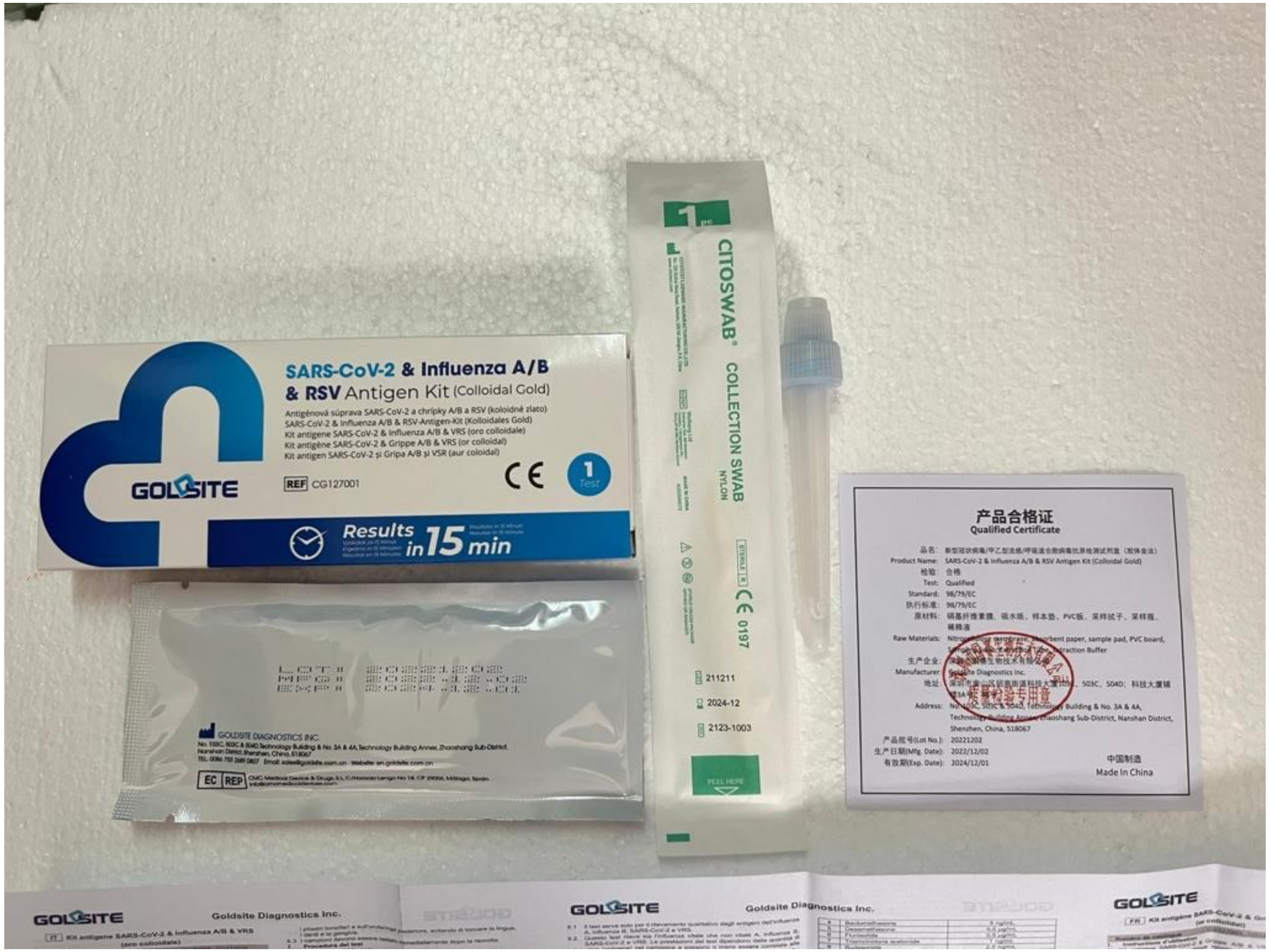

Appendix Figure 2: SARS-CoV-2 & Influenza A/B & RSV Antigen Kit (Colloidal Gold) (Goldsite Diagnostics Inc., Shenzhen, China)

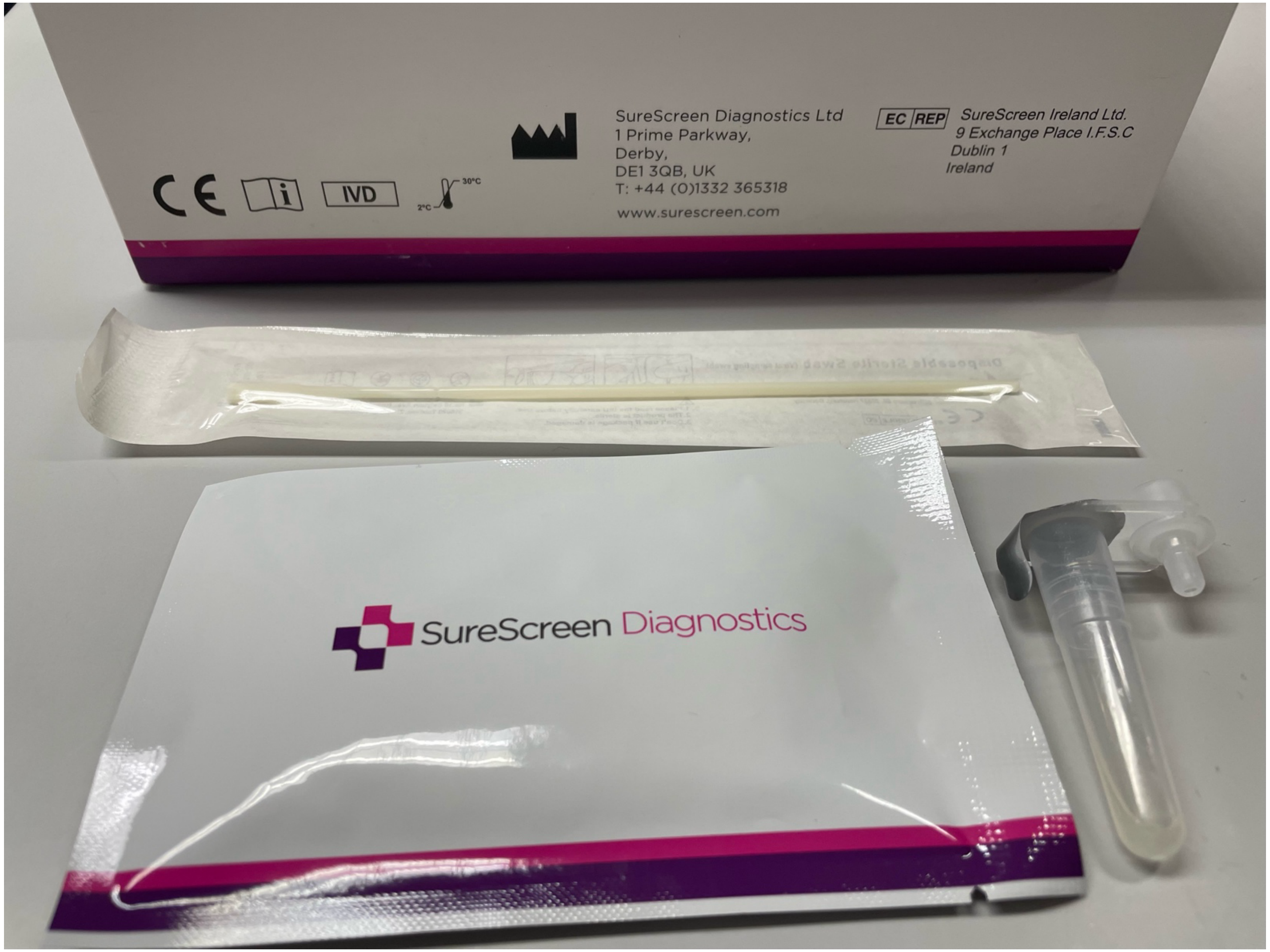

Appendix Figure 3: SARS-CoV-2 + Flu A&B Antigen Combo Rapid Test Cassette (Nasal Swab) (SureScreen Diagnostics Ltd, Derby, UK)

Appendix Table 3: Lateral flow test positivity in Influenza A, SARS-CoV-2 and RSV positives.

| | Multivariable regression model | | |
|---|---|---|---|
| | OR | 95% CI | p-value |
| **Influenza A positives (n=651)** | | | |
| Age | | | |
| 6 months to 5 years | Ref | - | - |
| 6 to 17 years | 0.91 | 0.38 to 2.01 | 0.826 |
| 18 to 49 years | 0.68 | 0.28 to 1.56 | 0.377 |
| 50 years and above | 0.22 | 0.09 to 0.55 | **0.001** |
| Sex | | | |
| Female | Ref | - | - |
| Male | 0.80 | 0.47 to 1.35 | 0.407 |
| Vaccination status | | | |
| Unvaccinated | Ref | - | - |
| Vaccinated | 0.98 | 0.52 to 1.90 | 0.948 |
| Symptom onset | | | |
| <=24 hours | Ref | - | - |
| >24 to 48 hours | 2.21 | 1.12 to 4.74 | **0.030** |
| >48 to 72 hours | 1.19 | 0.65 to 2.26 | 0.586 |
| **SARS-CoV-2 positives (n=171)** | | | |
| Age | | | |
| 6 months to 5 years | Ref | - | - |
| 6 to 17 years | 0.85 | 0.11 to 5.42 | 0.868 |
| 18 to 49 years | 1.82 | 0.26 to 10.76 | 0.518 |
| 50 years and above | 3.45 | 0.44 to 26.20 | 0.223 |
| Sex | | | |
| Female | Ref | - | - |
| Male | 0.96 | 0.33 to 2.85 | 0.935 |
| Vaccination status | | | |
| Unvaccinated | Ref | - | - |
| Vaccinated | 0.58 | 0.06 to 3.57 | 0.588 |
| Symptom onset | | | |
| <=24 hours | Ref | - | - |

| | | | |
|---|---|---|---|
| >24 to 48 hours | 0.80 | 0.26 to 2.71 | 0.695 |
| >48 to 72 hours | 1.07 | 0.25 to 7.39 | 0.936 |
| **RSV positives (n=75)** | | | |
| Age | | | |
| 6 months to 5 years | Ref | - | - |
| 6 to 17 years | 0.58 | 0.08 to 5.12 | 0.593 |
| 18 to 49 years | 0.27 | 0.02 to 2.86 | 0.256 |
| 50 years and above | -* | - | - |
| Sex | | | |
| Female | Ref | - | |
| Male | 0.72 | 0.09 to 4.45 | 0.725 |
| Symptom onset | | | |
| <=24 hours | Ref | - | - |
| >24 to 48 hours | 2.65 | 0.22 to 61.77 | 0.451 |
| >48 to 72 hours | 0.66 | 0.07 to 4.44 | 0.680 |

** All RSV positives in the older adults age group where successfully identified by lateral flow tests.*

*OR = Odds ratio*

*CI = confidence interval*

Appendix Table 4: Characteristics of the false positives reported by RAT compared to RT-PCR confirmed viral infection.

| | **Influenza A (n=138)** | **SARS-CoV-2 (n=30)** | **RSV (n=5)** |
|---|---|---|---|
| **Male** | 57 (41.3) | 15 (50.0) | 3 (60.0) |
| **Underling medical condition** | 4 (2.9) | 0 (0.0) | 0 (0.0) |
| **Age** | | | |
| 6 months to 5 years | 35 (25.4) | 10 (33.3) | 1 (20.0) |
| 6 to 11 years | 41 (29.7) | 3 (10.0) | 1 (20.0) |
| 12 to 17 years | 10 (7.2) | 1 (3.3) | 1 (20.0) |
| 18 to 49 years | 44 (31.9) | 13 (43.3) | 1 (20.0) |
| 50 to 64 years | 7 (5.1) | 2 (6.7) | 1 (20.0) |
| 65 years and above | 1 (0.7) | 1 (3.3) | 0 (0.0) |
| **RT-PCR confirmed infection** | | | |
| Influenza A | - | 3 (10.0) | 3 (60.0) |
| Influenza B | 0 (0.0) | 0 (0.0) | 0 (0.0) |
| SARS-CoV-2 | 8 (5.8) | - | 1 (20.0) |
| RSV | 5 (3.6) | 1 (3.3) | - |

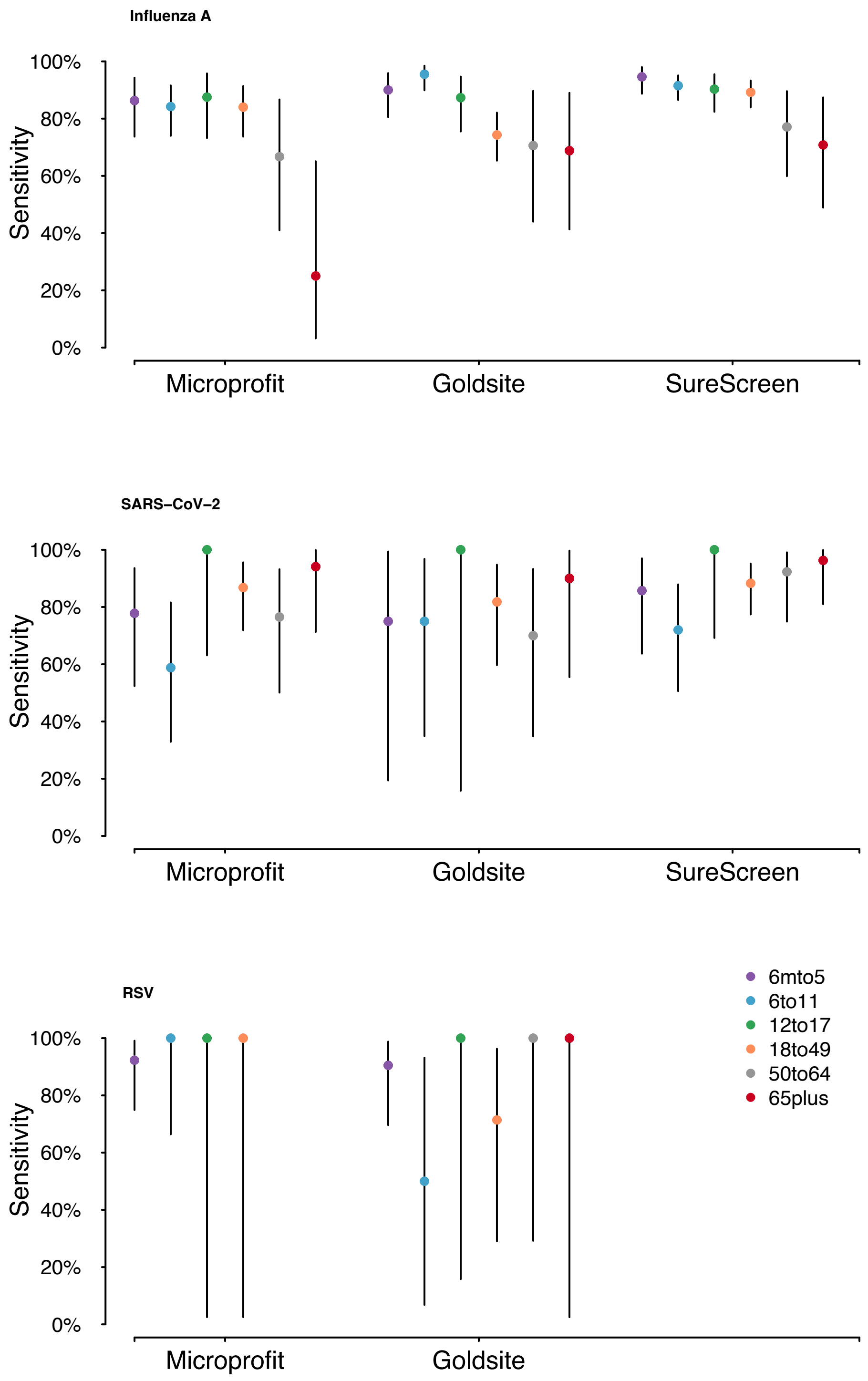

Appendix Figure 4: Test specific sensitivity with 95% confidence intervals by age group.
